# Optimal Control Strategies for Mitigating Antibiotic Resistance: Integrating Virus Dynamics for Enhanced Intervention Design

**DOI:** 10.1101/2024.12.07.24318622

**Authors:** Zainab Dere, N. G. Cogan, Bhargav R. Karamched

## Abstract

Given the global increase in antibiotic resistance, new effective strategies must be developed to treat bacteria that do not respond to first or second line antibiotics. One novel method uses bacterial phage therapy to control bacterial populations. Phage viruses replicate and infect bacterial cells and are regarded as the most prevalent biological agent on earth. This paper presents a comprehensive model capturing the dynamics of wild-type bacteria (*S*), antibiotic-resistant bacteria (*R*), and infective (I) strains, incorporating virus inclusion. Our model integrates biologically relevant parameters governing bacterial birth rates, death rates, and mutation probabilities and incorporates infection dynamics via contact with a virus. We employ an optimal control approach to study the influence of virus inclusion on bacterial population dynamics. Through numerical simulations, we establish insights into the stability of various system equilibria and bacterial population responses to varying infection rates. By examining the equilibria, we reveal the impact of virus inclusion on population trajectories, describe a medical intervention for antibiotic-resistant bacterial infections through the lense of optimal control theory, and discuss how to implement it in a clinical setting. Our findings underscore the necessity of considering virus inclusion in antibiotic resistance studies, shedding light on subtle yet influential dynamics in bacterial ecosystems.

## 1. Introduction

Bacteria are microorganisms that are composed of a single cell. They are widely distributed in nature and cause a range of infections and also play an essential role in human health [1]. Beneficial bacteria can be found in the gastrointestinal tract, where they support the digestive system and aid healthy development of the immune system. *Bifidobacteria* and *E. coli* are examples of healthy bacteria found in the intestine which break down complex carbohydrates and also improve gut health [2]. Mutations in the DNA of a bacteria may occur due to mistakes when bacterial cells undergo binary fission [1, 3] and can sometimes alter the functioning of its genes, leading to changes in their phenotype. These mutations facilitate the emergence of diversity within its population, which may improve the capacity of the bacteria to adapt to its changing environment [4–7]. When mutations occur in bacteria important for human health, harmful infections could arise. Typical treatment for such pathogenic bacterial infection involves antibiotics. However, some mutations render bacteria antibiotic-resistant, making it difficult to treat infections. Other mutations make bacteria more virulent or more easily transmissible [8–10].

Antibiotic resistance in bacteria is a growing problem in public health, as it is increasingly harder to treat infections caused by mutant antibiotic-resistant bacteria [3, 11]. The scourge of antibiotic-resistant genes among microbial pathogens poses a serious threat to the effectiveness of current antimicrobial treatments, particularly for severe bacterial infections leading to sepsis [12]. The progressive emergence of antimicrobial resistance (AMR) has been driven by the use of anti-infection treatments in humans, animals, and food production [10, 13]. This has been further compounded by the inadequacy of measures designed to curtail the infections [13]. According to the World Health Organization (WHO), antibiotic resistance has significant economic cost implications as a result of prolonged illnesses and longer hospital stays which could lead to death or disability [14]. Two central factors that drive antimicrobial resistance are the volume of antimicrobials used and the spread of resistant micro-organisms and genes encoding for resistance [15]. These factors can both be controlled through preventative measures. A central goal of infection bacteriology is thus to identify aspects of bacterial ecology to mitigate infection and develop novel treatments.

Mathematical models have been historically successful in capturing essential features of infective microbes to predict infection dynamics. These models have significantly advanced our understanding of infectious diseases, their transmission, and the development of new treatments. Jenner *et al*. [16] and Xavier *et al*. [17] both highlight the crucial role of innovative mathematical and computational modeling techniques in predicting disease outbreaks and designing containment strategies. Mathematical models have also been used to study the evolution of infectious microbes, including their adaptation to host immune systems and the emergence of new strains [18], which has helped to inform the development of new vaccines and drugs. Several authors have considered the mathematical modeling of antimicrobial resistance with different objectives. For example, Ibargüen-Mondragón *et al*. [19] proposed an ODE model for the concurrent acquisition of resistance to bactericidal and bacteriostatic antibiotics, where resistance is generated by specific changes in bacterial DNA sequence and plasmid transmission. The model showed that applying appropriate therapies and stimulating the immune system is the best way to eliminate progression to resistance for many bacterial infections. Alavez-Ramírez *et al*. [20] presented a model for the emergence of resistance of Mycobacterium tuberculosis bacteria to antibiotics to assess the efficiency of administering one or two drugs for controlling latent tuberculosis infection considering its dependence on strengths of the immune system. Regarding AMR control, a number of studies have established results for the optimal control for bacterial resistance. For managing bacterial populations with persister dynamics, Leenheer and Cogan [21] applied the amount of antimicrobial as the control variable to predict the optimal timing and duration of antibiotic treatment. Ibargüen-Mondragón *et al*. [22] formulated an optimal control problem to minimize the bacterial population with plasmid-mediated antibiotic resistance, considering the action of both antibiotic treatment and immune system to combat bacterial infections. Gutiérrez *et al*. [23] developed an approach for managing bacterial populations with persistent dynamics. They offer a completely automated, high-throughput approach that combines in-the-moment measurements with computer-controlled optogenetic manipulation of bacterial growth to perform precise and reliable compositional control of a two-strain *E. coli* community. From a deterministic perspective, we can single out other research focused on the acquisition of antibiotic resistance: the causal factors underlying bacterial resistance is given in [24], analysis of bacterial behavior in response to various antibiotic treatments in [25], [26], [27], optimum antibiotic use in [28], and modeling of the acquisition of resistance from external sources in [29]. Both deterministic and stochastic models have been used to study bacterial resistance mathematically. The interaction between antibiotic-sensitive and antibiotic-resistant bacteria is studied mathematically by Mena *et al*. [30]. They formulated an optimal control problem for an unperturbed and a perturbed system, where the control variable is prophylaxis. Merdan *et al*. [31] compared mathematical models of bacterial resistance under random conditions with a deterministic model including immune system response and antibiotic therapy. In [32], the authors applied a stochastic population model to investigate the effect of resistance, persistence, and hyper-mutation on antibiotic treatment failure and found that the relative impact of these factors depends on the antibiotic concentration and the infection time scale.

In this paper, we will focus on the use of viruses to mitigate bacterial infection. Roach and Donovan [33] explored the therapeutic applications of bacteriophage-derived proteins, such as endolysins and peptidoglycan hydrolases, in animal models of bacterial infection. The potential of targeting bacterial virulence and the use of bacteriophage-based approaches was highlighted in [34–36], as alternative strategies to combat antibiotic resistance and control bacterial infections. Mekalanos *et al*. [37] explored how bacteriophages can influence the dynamics of cholera outbreaks. The research showed that lytic bacteriophages, which specifically target and destroy virulent strains of cholera-causing bacteria (*Vibrio cholerae*), can significantly reduce the severity of cholera outbreaks. Furthermore, in their study on bacteriophage-resistant and bacteriophage-sensitive bacteria, Han and Smith [38] explored the population dynamics within a chemostat. They found that while resistant bacteria may survive phage attacks, they are less efficient at competing for nutrients compared to sensitive bacteria. This trade-off significantly influences their population dynamics and persistence.

Mathematical modeling has been instrumental in understanding the dynamics and control of viral infections, including the use of viruses to control bacterial infections [39]. Clifton *et al*. [40] demonstrated that antibiotic-induced proviruses can play a role in controlling bacterial populations, expanding the understanding of phage-antibiotic synergy. Bacteriophages, viruses that only replicate in and infect bacterial cells, are regarded as the most prevalent biological agent on earth and are found everywhere in the environment. Styles *et al*. [41] emphasized the need for realistic mathematical models to improve the understanding of bacteriophage, bacteria, and eukaryotic interactions, which is crucial for the development of phage therapy. These studies collectively underscore the importance of mathematical modeling in exploring the use of viruses to control bacterial infections. One challenge with phage therapy is that phages, like other pathogens, are construed as “outsiders” by the human immune system and are eliminated in due time [42, 43]. Thus, if phage therapy is unsuccessful once, the same phage therapy cannot be used to treat a given infected individual. Using virotherapy to completely eliminate a given infection is possible but with a low chance of success. However, this can be circumvented by using phages in combination with other antibacterial agents, including other phages [43, 44].

Here, we hope to explore phage therapy with a mathematical model in a different capacity. Rather than completely eliminating an infection, we seek to understand if introducing virus-infected bacteria can help mitigate an infection indefinitely. We borrow from the concept in ecology known as apparent competition, wherein the presence of multiple prey with a common predator prevents any single prey from being eliminated [45]. If we consider wild-type bacteria and antibiotic-resistant bacteria as prey for a bacteriophage, the theory of apparent competition stipulates that the mutant bacteria will not outgrow the wild-type bacteria. Thus, infection may not be completely removed, but it can be controlled. In this paper, we describe the dynamics of the interactions of antibiotic-sensitive and antibiotic-resistant bacteria with a user-controlled viral infection. To this end, we formulate a mathematical model that consists of a nonlinear system of three ordinary differential equations. These equations describe the interaction between populations of bacteria sensitive to and resistant to antibiotics, along with the viral infection. With the goal of minimizing the population of antibiotic resistant bacteria, we formulate an optimal control problem where the control variable is a proxy for introducing virus-infected bacteria near the region of bacterial infection. The infection is user-controlled as a mechanism to mitigate undesirable antibiotic resistant bacteria.

The study introduces a novel framework for understanding and optimizing control interventions in bacterial ecosystems. By integrating optimal control techniques, dynamically exploring control strategies, quantitatively assessing control impact, and elucidating long-term system behavior, we provide a holistic approach to tackling antibiotic resistance and advancing our understanding of complex ecological dynamics with the goal of reducing the antibiotic resistant population. Our work lays the foundation for progressing treatment of antibiotic-resistant bacterial infection.

## 2. Model Formulation

Our model consists of three different strains of bacteria: wild-type (S), antibiotic-resistant (R), and the infective (I) strains. It describes the interactions between them (see Figure 1). Analyzing this model will provide insight into which biophysical parameters are key to affecting model output. Let *S*(*t*), *R*(*t*) and *I*(*t*) be the densities (*S, R, I >* 0) of wild-type antibiotic bacteria, mutant antibiotic resistant bacteria, and virus-infected bacteria, respectively. The interactions between them can be represented by the following model:

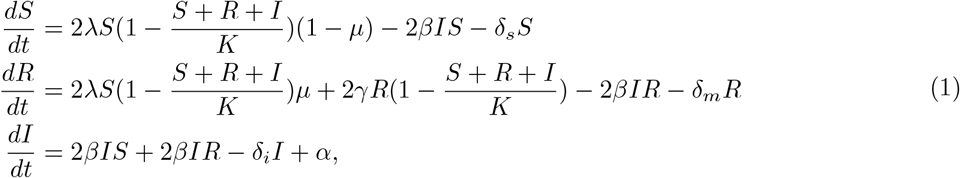

where *µ* is the probability that a wild-type bacterial division results in an antibiotic-resistant mutation. We assume *µ* depends on *R*: As *R* increases, the likelihood of horizontal gene transfer increases, yielding more mutants. We therefore take *µ* to be an increasing function of *R* that saturates as *R →∞*. We represent this with a Hill function:

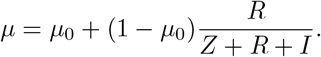

**Figure 1.**
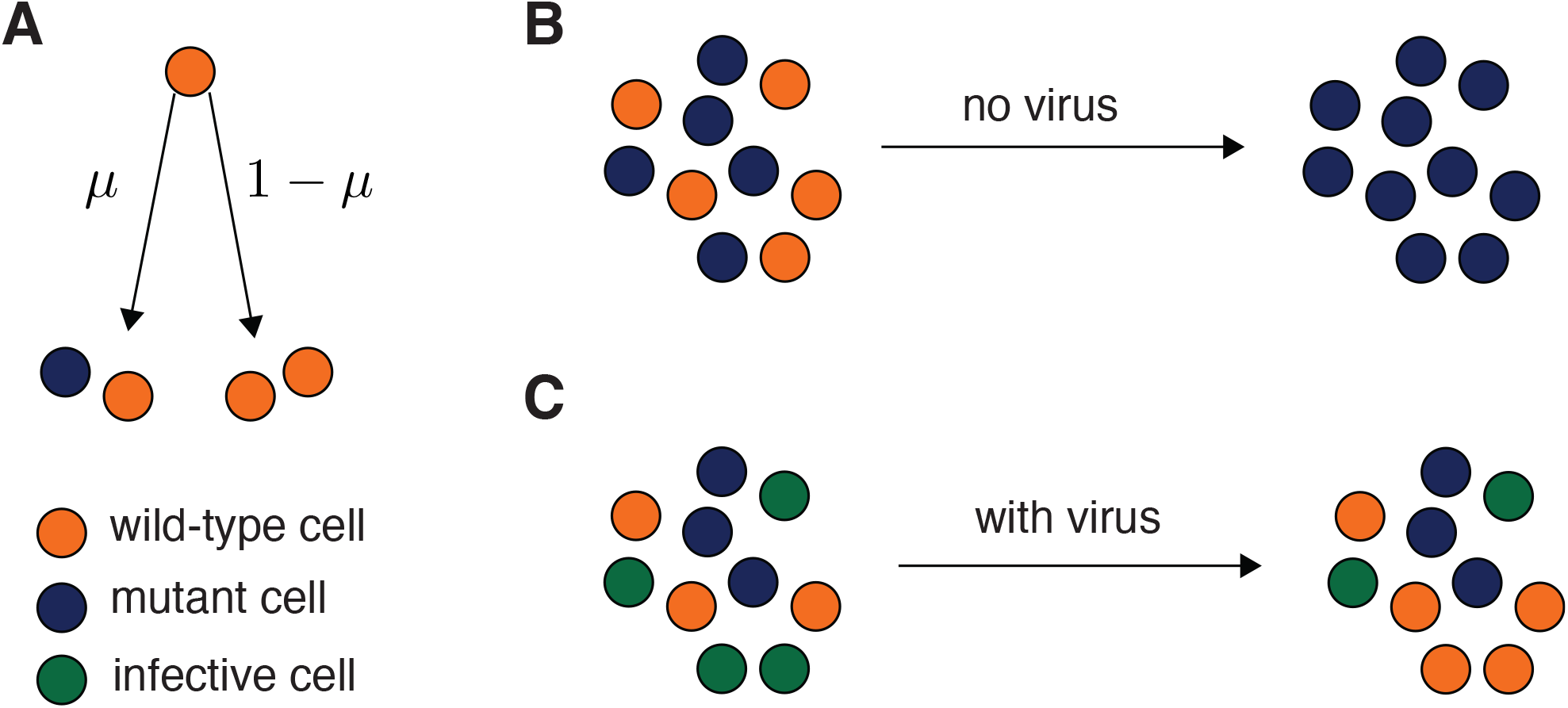
Schematic of our model. (A) Wild-type cells undergo fission and with a probability *µ* produce mutants that are antibiotic-resistant. (B)If mutants arise, then they will eventually dominate the population. This is undesirable. (C) We hypothesize that adding viral infection promotes the ability for coexistence between the wild-type and mutant strains.

We assume a basal mutation probability *µ*_0_, which is the inherent likelihood of a mutation occurring independent of the number of the antibiotic-resistant bacteria. The parameter *Z* is the half-activation concentration. The birth rate and death rate of wild-type bacteria are *λ* and *δ*_*s*_^1^, respectively, and we assume a logistic growth described by *S*(1 *− S − R − I*) with carrying capacity *K*. For the mutant strain, the birth rate and death rate are characterized by *γ* and *δ*_*m*_, respectively. Mutation of wild-type bacteria leading to new resistant bacteria is represented with *S*(1 *– S − R − I*)*µ*. Our model forgoes explicit virus dynamics and models infection via contact with infected bacteria. The rate at which we inject virus-infected bacteria into the system is described by *α*, and *β* quantitates the infectivity of the viral infection. We assume infection is a stronger contributor to infective cell growth than division—our model thus does not include a term for infective cell birth [46, 47]. Including such a term does not qualitatively affect our results. The death rate of infective cells is characterized by *δ*_*i*_. Here, we assume all the model parameters *λ, γ, β, δ*_*s*_, *δ*_*m*_, *δ*_*i*_ are positive and that all the initial conditions of model system (1) satisfy *S*(0), *R*(0), *I*(0) *≥* 0. For simplicity, we set *K* = 1 for the rest of the paper. Other parameter values are given in Table 1. Unless otherwise noted, these are the values used throughout the paper.

**Table 1.**
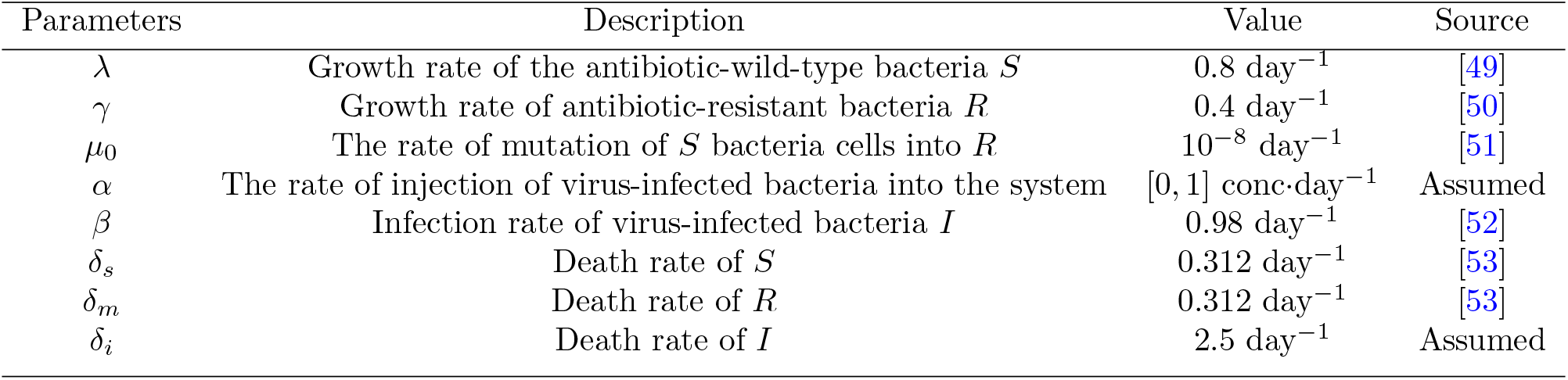
Description of parameters used.

| Parameters | Description | Value | Source |
| --- | --- | --- | --- |
| $\lambda$ | Growth rate of the antibiotic-wild-type bacteria $S$ | $0.8 \text{ day}^{-1}$ | [49] |
| $\gamma$ | Growth rate of antibiotic-resistant bacteria $R$ | $0.4 \text{ day}^{-1}$ | [50] |
| $\mu_0$ | The rate of mutation of $S$ bacteria cells into $R$ | $10^{-8} \text{ day}^{-1}$ | [51] |
| $\alpha$ | The rate of injection of virus-infected bacteria into the system | $[0, 1] \text{ conc} \cdot \text{day}^{-1}$ | Assumed |
| $\beta$ | Infection rate of virus-infected bacteria $I$ | $0.98 \text{ day}^{-1}$ | [52] |
| $\delta_s$ | Death rate of $S$ | $0.312 \text{ day}^{-1}$ | [53] |
| $\delta_m$ | Death rate of $R$ | $0.312 \text{ day}^{-1}$ | [53] |
| $\delta_i$ | Death rate of $I$ | $2.5 \text{ day}^{-1}$ | Assumed |

We note that Eq. (1) can be derived systematically from a lattice-based microscopic spatial stochastic model of bacterial dynamics. In such a model, each lattice site is occupied by a wild-type, mutant, or infective bacterial cell or is vacant. Division and infection events characterize the reactions that describe the stochastic evolution of the microscopic configurations of the lattice. By invoking a mean field approximation and a macroscopic limit, one can derive Eq. (1) [7, 48]. We will explore this stochastic lattice model in detail as a subject of future work.

To understand what biophysical parameters promote reaching desirable stationary states, we next compute the equilibria of Eqs. (1) and determine their stability in the absence of the control *α* (*α* = 0). Then, using *α* as our control variable, we will apply optimal control to the system to determine how to tune *α* in time to minimize the antibiotic resistant bacteria population while maintaining the wild-type bacteria population.

## 3. Model Analysis

This section explores the dynamics of bacterial populations as the infection rate increases. It includes simulations of the bacteria population under two conditions: one without any control measures and no initial infection, and another with an increased infection rate and an initial infection present. These analyses highlight how varying infection rates and initial conditions affect population trajectories, providing insights into the effectiveness of infection control strategies.

To examine these various circumstances, we compute the equilibria of Eq. (1) and determine their respective stability.

### 3.1. Linear Stability Analysis

We find the equilibria of system (1) by setting the temporal derivatives on the left hand side to zero and solving the resulting algebraic equations.

*No virus case*. In the absence of virus and control (*β* = 0, *α* = 0, *I*(0) = 0), Eq. (1) is reduced to a planar system and two equilibria manifest. One equilibrium (*S* = 0, *R* = 0) represents an extinction scenario where no bacteria are present. The eigenvalues of the corresponding linearization are

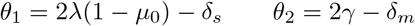

The instability of this state for our parameter values, indicated by the positive eigenvalues in the linearization (see Table 2), suggests that any small perturbation away from this equilibrium will cause the population to move away from extinction towards an alternative equilibrium. Consequently, the bacteria strains rapidly proliferate, but a significant concern arises if the antibiotic-resistant population begins to grow faster than the wild-type population. This scenario would result in the resistant strains becoming predominant, posing a significant challenge for treatment and control of infections.

**Table 2.**
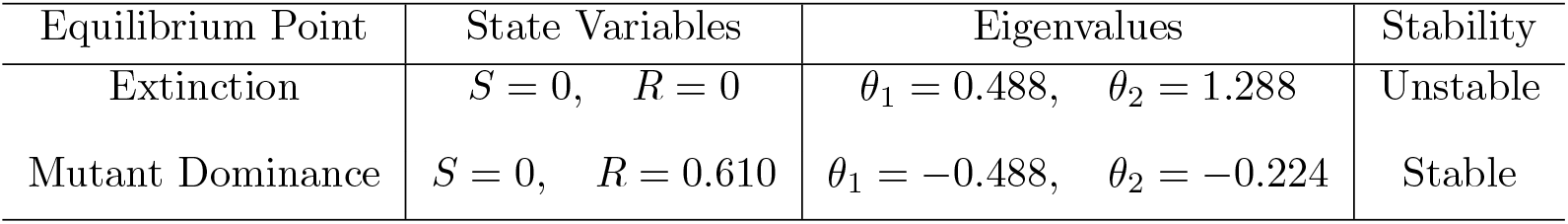
Steady states and stability analysis when *β* =0, *α* =0 and no infectives present with parameter values.

We observed this exact occurrence manifest as the other equilibrium point from Eq. (1). The extermination of wild-type bacteria by the resistant strain 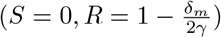 occurs as a stable equilibrium (see Table 2, Figure 2A), with eigenvalues corresponding to the linearization

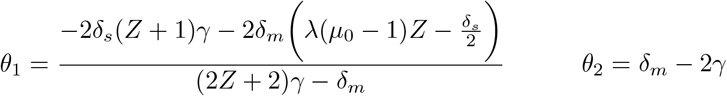

being negative for our parameter values. Thus, any small perturbation away this state will return to the equilibrium, which characterizes the dominance of the antibiotic-resistant population. This outcome is undesirable because it implies that the antibiotic-resistant bacteria have become the predominant strain, making it difficult to control infections.

**Figure 2.**
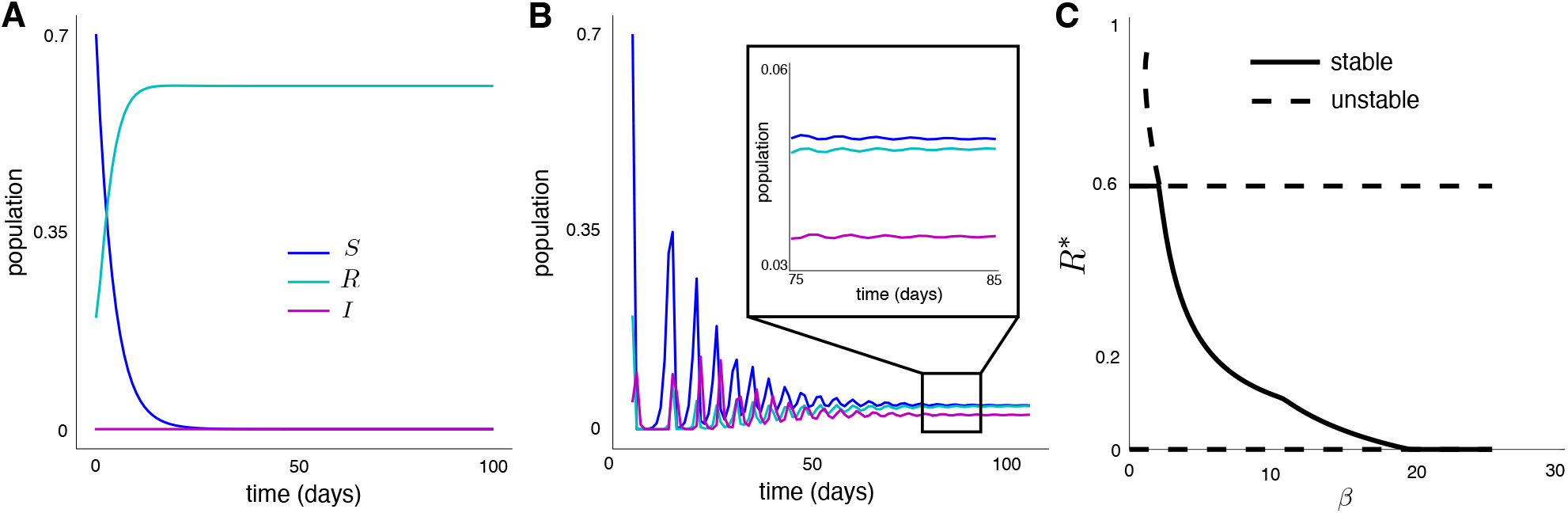
Simulation results of Eq. (1): (A) bacteria population in the absence of virus-infected bacteria *α* = 0, *β* = 0 and *I*(0) = 0, (B) bacteria population in the presence of virus-infected bacteria *α* = 0, *β* = 15 and *I*(0) *>* 0, and (C) a bifurcation diagram showing the impact of virus infectivity on equilibrium mutant population.

The proliferation of resistant bacteria highlights the critical need for the development of new antimicrobial treatments and the implementation of rigorous infection control measures. Addressing this issue is essential to prevent the rise of untreatable bacterial infections and safeguard public health. These findings underscore the urgency of implementing optimal control measures to prevent the growth of resistant strains and to manage existing populations effectively.

*Adding virus-infected bacteria*. To ascertain if imputing a viral intervention in system dynamics allows for resistant population mitigation, we next introduce a virus infected population into our system and determine the stability of various equilibria from Eq. (1). Following a similar process applied to the no virus case summarized in Table 2, we obtain the equilibria and the eigenvalues for the model when the virus infected population is added. In this scenario, analytical results for the equilibria and eigenvalues are obtainable, but are extremely complicated to parse. As such, we present numerical results for our parameter set.

Four equilibria result: extinction, mutant dominance, mutant-infective coexistence, and coexistence (see Table 3):

1. Extinction: The linearization around the extinction scenario (*S* = 0, *R* = 0, *I* = 0) yields eigenvalues with positive real part. Thus, it is unstable. This suggests that both bacterial strains and infective agents could proliferate rapidly when pushed away from extinction (unless the dynamics are confined to a 1D stable manifold, which we consider unlikely). There are two possibilities that emerge from this instability: (1) The mutants again dominate or (2) the infectives facilitate coexistence.
2. Mutant Dominance: The possibility of strict mutant dominance is nullified in the presence of virus because the mutant dominant equilibrium is unstable. This instability suggests that the introduction of the virus can alter the system dynamics, leading to the growth of the bacteria strains over time. Although there exists a planar subspace of phase space wherein contraction to the mutant-dominant equilibrium occurs, we do not consider this as a generic perturbation away from this equilibrium. This highlights the impact of the virus in mitigating the dominance of antibiotic resistance in the bacterial populations.
3. Mutant-Infectiv0065 Coexistence: This equilibrium represents a case where mutant (resistant) bacterial strains coexist with infective bacteria and the wild type bacteria have gone extinct. The eigenvalues indicate instability, suggesting that small disturbances will cause the populations to deviate from this delicate balance. The instability of this steady state will support the growth of the wild-type bacteria population. The instability implies that both resistant and infective bacterial populations could grow faster than the wild-type strains, potentially leading to their dominance. This outcome is undesirable, necessitating the implementation of effective control measures to prevent the unchecked proliferation of resistant and infective bacterial strains.
4. Coexistence: In this scenario, all bacterial strains coexist. Importantly, this equilibrium is stable as indicated by the negative real parts of all eigenvalues in the linearization. Furthermore, in this case with our parameters, the model predicts that the wild-type strain outnumbers the mutant, which is desirable (see Figure 2B). This ensures a level of control and mitigation of the risks associated with bacterial coexistence and resistance emergence.

**Table 3.** Equilibria and stability when *β >* 0, *α* = 0, and *I >* 0. Here, 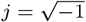.

| Equilibrium Point | State Variables | Eigenvalues | Stability |
| --- | --- | --- | --- |
| Extinction | $S = 0, R = 0, I = 0$ | $\theta_1 = 0.488$<br>$\theta_2 = 1.288$<br>$\theta_3 = -2.500$ | Unstable |
| Mutant Dominance | $S = 0, R = 0.610, I = 0.0$ | $\theta_1 = -0.488$<br>$\theta_2 = -0.224$<br>$\theta_3 = 15.800$ | Unstable |
| Mutant Infective Coexistence | $S = 0, R = 0.083, I = 0.014$ | $\theta_1 = 0.111$<br>$\theta_2 = -0.0333 + 1.0258j$<br>$\theta_3 = -0.0333 - 1.0258j$ | Unstable |
| Coexistence | $S = 0.042, R = 0.041, I = 0.025$ | $\theta_1 = -0.0617$<br>$\theta_2 = -0.0639 + 1.4389j$<br>$\theta_3 = -0.0639 - 1.4389j$ | Stable |

Thus, we have established that the presence of virus mitigates the mutant (resistant) strain of bacteria. To further embellish this point, we show a bifurcation diagram in Figure 2C depicting how the equilibrium resistant population decreases as a function of virus infectivity (*β*). For low infectivities, the mutant strain is able to maintain its dominance. However, at a critical infectivity, a transcritical-like bifurcation occurs and stabilizes the coexistence state. In principle, introducing a highly infective virus will completely exterminate the mutants. This is desirable, but a high infectivity will eventually eliminate all bacterial strains, which is not desirable.

Although simply introducing virus-infected bacteria helps mitigate resistant bacterial infection, the persistence of resistant strains at levels similar to wild-type strains is problematic. Furthermore, the stable equilibrium suggests coexistence, but the vast majority of the compartment is vacant (1 *− S*^*\**^ *− R*^*\**^ *− I*^*\**^ = 0.892). Thus we must determine if it is possible to attain a situation where the wild-type strain outnumbers the resistant strain significantly and populates much of the compartment. To that end, we implement optimal control theory on Eq. (1).

### 3.2. Optimal Control

The integration of optimal control techniques in studying bacterial dynamics, particularly in the context of antibiotic resistance and virus inclusion, is motivated by the urgent need for effective strategies to combat bacterial infections. Optimal control methods offer a powerful framework for designing and implementing interventions that optimize the use of available resources, such as antibiotics and vaccines, to minimize the emergence and transmission of resistant strains while preserving the efficacy of existing treatments. Recent studies have highlighted the potential of optimal control in guiding decision-making processes and informing policy interventions aimed at controlling antibiotic resistance [54–56]. By incorporating virus inclusion into the model, our study further extends this framework to elucidate the complex interplay between bacteria and viruses and to explore novel avenues for combating antibiotic resistance.

For the purposes of mitigating resistant bacteria, consider the objective function

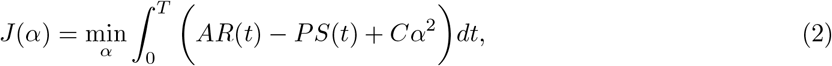

subject to Eq. (1) with

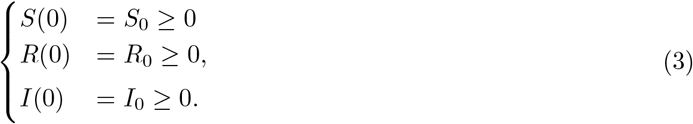

The constants *A, P* and *C* are weights applied on *S*(*t*), *R*(*t*) and the control variable *α* (the virus infected bacteria infusion rate), respectively. We employ a standard quadratic term for the control variable *α* in equation (2) following [57]^2^. The goal is to minimize the objective functional *J* (*α*) in order to find the optimal values of *α* such that the antibiotic-susceptible bacteria population *S* is maximized while the antibiotic-resistant bacteria population *R* is minimized. The objective of minimizing the infected population and the cost of control can be achieved through proper implementation of the control over a time interval given by [0, *T*]. Therefore, we seek an optimal control *α*^*^ such that

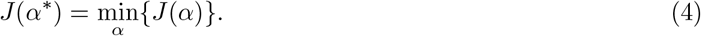

The necessary conditions for the existence of an optimal solution come from Pontryagin’s Maximum Principle [57]. This principle converts Eqs. (2)–(3) into a problem of minimizing the associated Hamiltonian *H* pointwise with respect to *α*. We define the Hamiltonian for this problem by

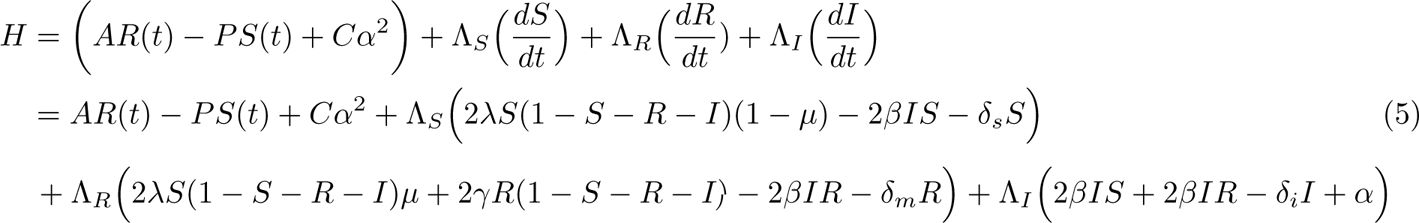

where Λ_*S*_(*t*), Λ_*R*_(*t*), and Λ_*I*_ (*t*) are the corresponding adjoint or co-state variables to be determined by applying Pontryagin’s Maximum Principle with the following transversality conditions

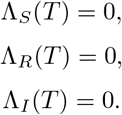

Using the optimality condition 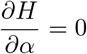, we solve for the optimal injection rate of the virus infected bacteria α.

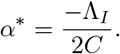

We solve the optimal control problem employing the forward-backward sweep method [57].

## Results

We implemented the optimal control over a period of *T* = 100 days and observed the resulting dynamics of the bacterial population state variables. Figure 3A shows the optimal trajectory of the control variable *α* to obtain our desired results. It predominantly maintains a constant value with sharp declines near the start and end of the time interval of interest. The effect of this control upon the bacterial strain populations is shown in Figures 3B-D. For comparison, we also present the analogous bacterial populations in the absence of control. Figure 3A shows that the wild-type bacteria population is significantly higher with control present than without it. Conversely, the antibiotic-resistant bacteria population *R* (see Figure 3B) decreased drastically in the presence of control. This finding underscores the intricate relationships between viruses and bacteria, as well as the potential for exploiting viral therapy to combat antibiotic resistance. The population dynamics when optimal control strategies are applied and its effect on the bacteria population provide insights into the dynamics of wild-type and antibiotic-resistant bacteria populations under different control scenarios, facilitating the evaluation and optimization of control strategies to manage antibiotic resistance effectively.

**Figure 3.**
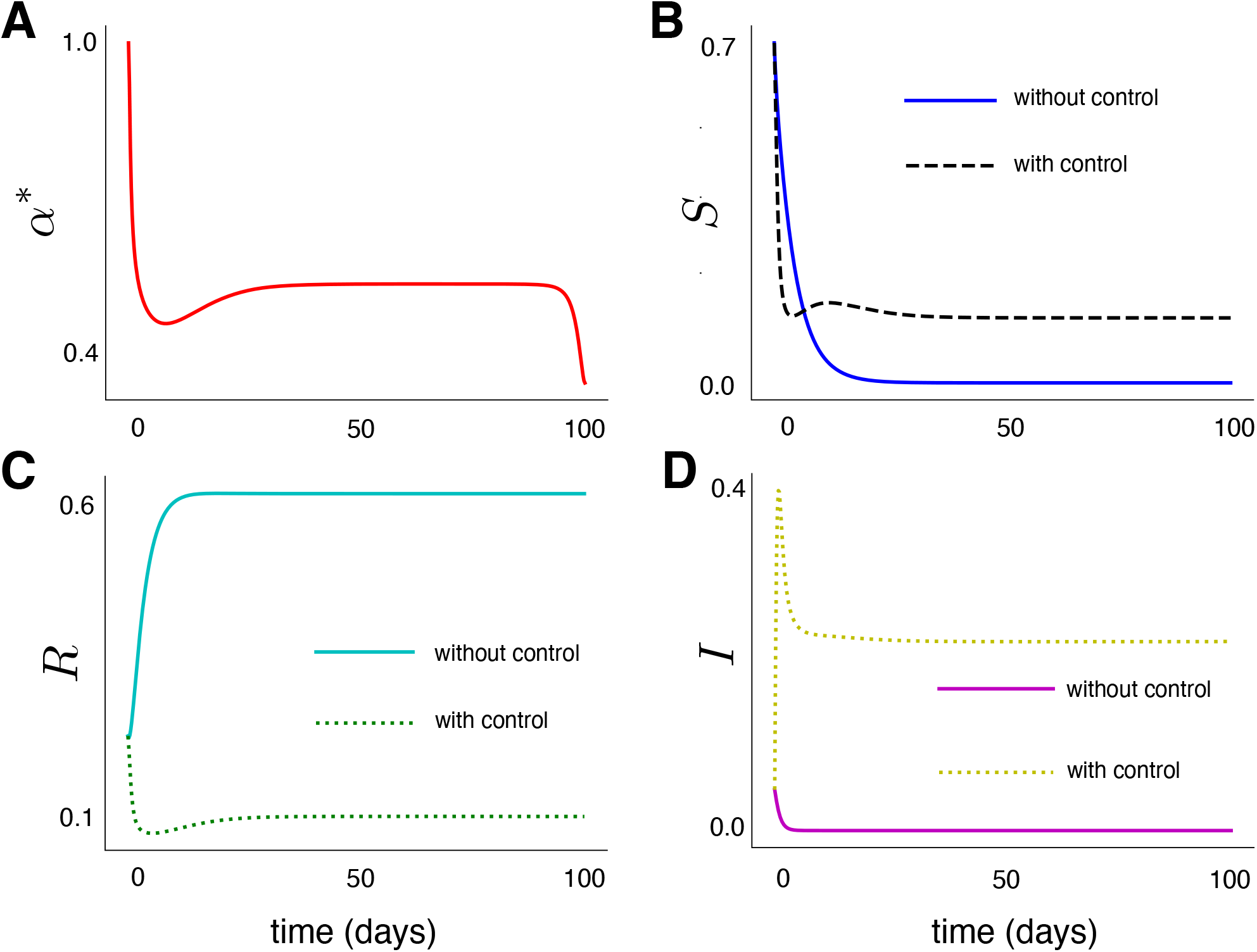
Simulation results of Eq. (1) with optimal control obtained from solving Eq. (4). (A) the optimal profile for the control variable *α*, (B) the corresponding wild type bacteria population *S*, (C) the corresponding antibiotic-resistant bacteria population *R*, and (D) the corresponding virus infected bacteria population *I*.

*A clinically plausible implementation*. The following results are motivated by the difficulty of clinically realizing the exact control profile in Figure 3A for the rate of infusion of the virus into the system. Although the optimal control profile facilitated the onset of a state where wild-type bacteria significantly dominate mutant strains, implementing such a tightly regulated control is not feasible clinically. However, because the control profile is predominantly constant, we seek to determine a constant injection rate 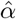 such that

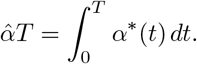

That is, we determine a constant injection rate 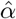 such that the total virus injected over the time interval [0, *T*] is equal to the total amount injected with the optimal control profile. This yields the simple equation for 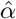 :

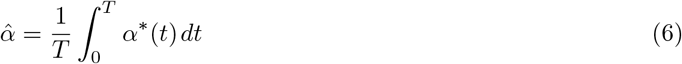

This constant rate of virus injection is applied into our system to see the effect on the dynamics of *S* and *R* and *I*. Success in reducing resistant bacteria with constant injection rate imputes stability in intervention measures while offering predictability.

Figure 4A-B illustrate the changes in the populations of *S* and *R*, respectively, reflecting the effect of the constant rate of viral infusion (Figure 4A inset) on the population in the bacterial community. The resulting dynamics of the bacterial strain populations are similar to Figure 3, indicating that system dynamics are not overtly sensitive to the control profile. In this clinically feasible injection protocol, desirable reduction in mutant bacterial population is achieved when compared with the absence of control case.

**Figure 4.**
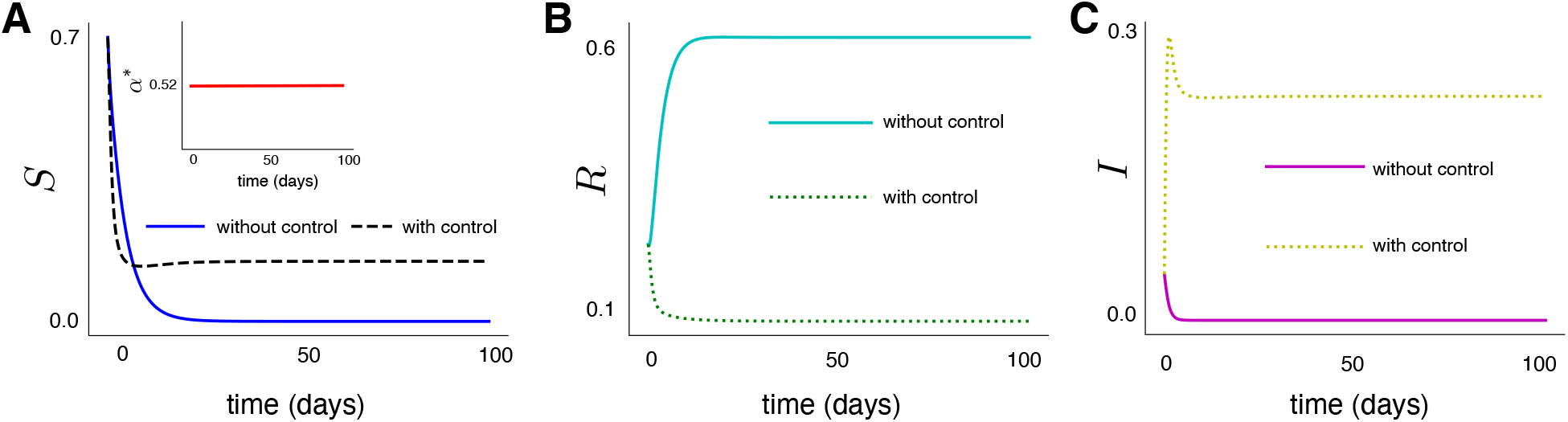
Simulation results from Eq. (1) with a constant virus infusion rate 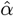 as obtained from Eq. (6) (A) Wild type population in response to the constant control profile. Inset: the constant control profile. (B) Antibiotic-resistant bacteria population *R* and (C) Virus infected bacteria population *I*.

How far from optimality is the constant control profile implementation? In Table 4 we compare the objective functional *J* for the different scenarios of *α* (Optimal vs Constant). The objective functional value for 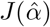 is reasonably close to *J* (*α*^*^), with approximately a 4% difference in value. Thus, implementing a constant viral infusion is a clinically feasible protocol that is near optimality for mitigating antibiotic-resistant mutant bacterial infection.

**Table 4.** Comparison of the the objective functional of the optimal and the constant control profile.

| Control Profile | Objective Functional |
| --- | --- |
| Optimal: $\alpha^* = [0, 1]$ | $J(\alpha^*) = 87.6903$ |
| Constant: $\hat{\alpha} = 0.5200$ | $J(\hat{\alpha}) = 90.3541$ |

## 4. Conclusion

This study focused on modeling the interplay between wild type bacteria, mutant antibiotic resistant bacteria, and virus-infected bacteria populations, with the specific aim of understanding how antibiotic-resistant bacterial infections may be treated or controlled in the presence of a viral infection. We found that the simple introduction of a virus facilitates the desirable outcome of wild-type bacteria outcompeting mutant resistant bacteria. However, the presence of virus vastly diminished the total bacterial population. This motivated the implementation of optimal control upon the viral infusion rate. In the presence of control, wild-type bacteria vastly overcome mutant bacteria. Although the optimal infusion profile is not realistically realizable in a clinical setting, we showed that using a constant profile approximation of the optimal infusion rate is sufficient to reduce resistants and maximize wild type bacteria.

Importantly, we did *not* seek to *eliminate* resistant bacteria completely. Our goal was simply to mitigate resistant bacteria population. This removed the need for unnecessarily strong interventions—such as the ones that facilitated onset of antibiotic-resistant mutant bacterial strains. Rather, we modified a standard dynamical system of bacterial dynamics in infections and allowed the intrinsic dynamics to result in desirable outcome of diminished mutant population.

The findings of this study have significant implications for curbing antibiotic resistance. Our strategy not only reduces the population of resistant bacteria but also maximizes the presence of wild-type bacteria, which are more susceptible to antibiotics. This dual benefit highlights the potential of using viral infections as a complementary tool in the fight against antibiotic resistance. Moreover, our study underscores the importance of considering the broader ecological and evolutionary dynamics in the design of treatment strategies. By not aiming to eliminate resistant bacteria completely, we avoid the selective pressures that often lead to the emergence of even more resistant strains. Instead, our approach seeks to balance the bacterial ecosystem, promoting a more sustainable and long-term solution to antibiotic resistance.

There are a number of issues to explore in future work. First, our work here demonstrates that viral infection could be a measure used to mitigate mutant resistant bacterial infection, but it clearly overlooks aspects of bacterial infection. For example, our model overlooks the effect of including spatial dynamics. It would be interesting to use dispersion theory to characterize the biophysical parameters that affect viral infection of resistant bacteria and to inform optimal control in space. Another important question in spatial systems is how do we incorporate viral dynamics? We can simply proceed as we did in this manuscript and track infected bacterial cells or we could explicitly model viral dynamics and their infection process of bacteria, leading to a complex multiscale modeling paradigm. Finally, as we noted in the body of manuscript, Eq. (1) can be derived systematically from a stochastic lattice model. It would be interesting to implement stochastic optimal control theory and see what wrinkles including noise brings to the dynamics.

## 5. Appendix

The adjoint equations for the optimal control are given by

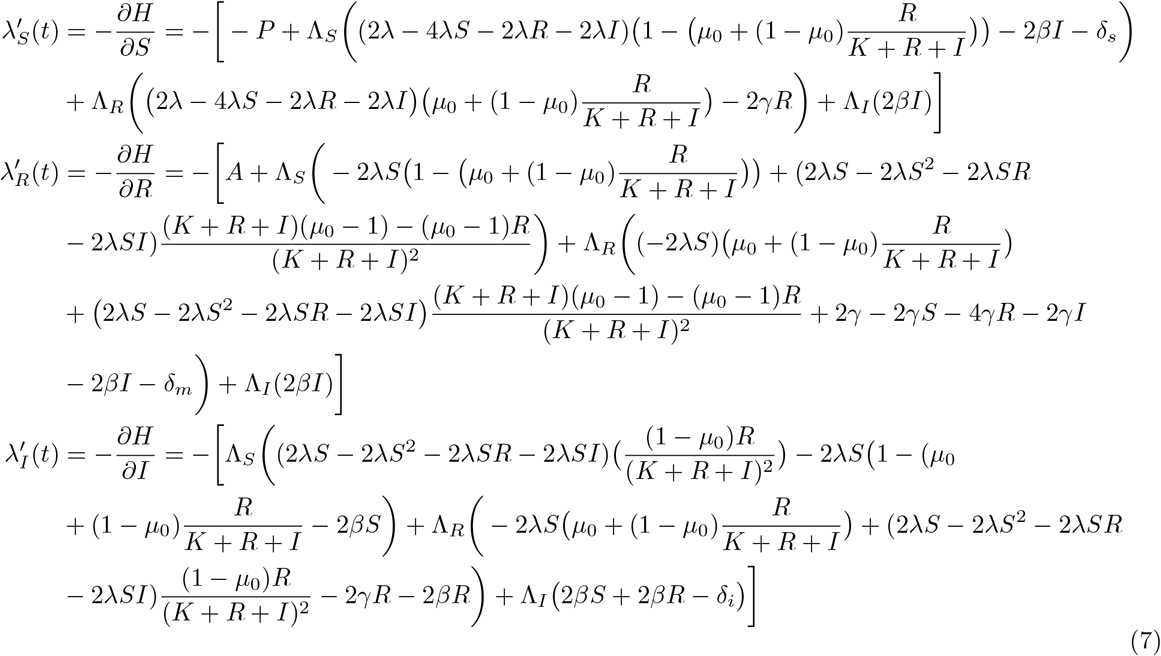

## Data Availability

All data produced in the present work are contained in the manuscript

## Footnotes

1 The death rates are not explicitly required since the logistic term accounts for death as well. We include this term to make the model amenable for future modifications, such as including antibiotic treatment.

2 Using a quadratic term facilitates onset of continuous control, whereas employing a linear term results in discontinuous bang-bang control

